# A review of the application of digital phenotyping in predicting peripartum depressive symptoms

**DOI:** 10.1101/2025.09.17.25335179

**Authors:** Boglarka Z Kovacs, Sascha Schweitzer, Fotios C Papadopoulos, Annette Bauer, Alkistis Skalkidou, Hsing-Fen Tu

**Affiliations:** Erasmus University Medical Center, Erasmus MC, Rotterdam, Netherlands; ESB Business School, Reutlingen University, Reutlingen, Germany; Faculty of Law and Economics, University of Bayreuth, Bayreuth, Germany; Department of Medical Sciences, Psychiatry, Uppsala University, Uppsala, Sweden; Care Policy and Evaluation Centre, the London School of Economics and Political Science, London, UK; Department of Women’s and Children’s Health, Uppsala University, Uppsala, Sweden; Department of Psychology, Uppsala University, Uppsala, Sweden; Department of Applied Educational Science, Umeå University, Sweden

## Abstract

Peripartum depression (PPD) affects 12 to 25% pregnant women worldwide, yet screening often misses real-time symptom changes. Digital phenotyping (DP) offers a promising support, using data like text entries or sleep tracking to detect PPD. This review (PROSPERO: CRD42023461325) evaluated 14 studies, highlighting the substantial potential of personal history and semi-random ecological-momentary data. Future work should focus on improving models and advancing their translation into clinical settings for broader impact.

## Introduction

Peripartum depression (PPD) affects nearly one in four to eight pregnant women^1,2^, adversely influencing parental health, mother-infant interactions^3^, and child development^4^. There is growing interest in incorporating *digital phenotyping* (DP) data, which provides moment-to-moment information (e.g., movement patterns) in natural settings^5^ and continuous mental state monitoring^6^, potentially enhancing early PPD identification and prediction. Most screening methods are restricted to one or two time points and often miss symptom fluctuations during the perinatal period^7,8^.

Accurate prediction, early identification, and monitoring of PPD are essential to personalized care^9,10^. Early screening programs is recommended by major health institutions, including the United States Preventive Services Task Force^11^, the American College of Obstetricians and Gynecologists^12^, and the Australian National Guidelines^13^, and the United Kingdom National Institute for Health and Care Excellence^14^. Yet, only 30.8% of women with PPD are diagnosed, and of those, merely 15.8% receive treatment^15^. Furthermore, common screening programs often rely on questionnaires at specific time points, missing many cases.

Although biological factors like hypothalamic-pituitary-adrenal dysregulation, and inflammatory markers have been linked with PPD^16^, the absence of consistent biomarkers complicates diagnosis^17^. Research also indicates that a lack of precise screening tools for early identification or prediction of women at risk for PPD and potential subtypes^18,19^, exacerbates these challenges. While psychosocial and environmental risk factors, such as psychiatric history, traums, poor socioeconomic status, poor social support^20,21^, and vulnerable personality^22^, have been identified, integrating DP and mobile health may provide a scalable solution for improving early PPD prediction^23^.

DP data offers low-cost, high-dimensional, and multi-modal continuous measurements, with potential to refine prediction models and facilitate timely, tailored interventions. However, the impact of differrnt DP types on enhancing prediction accuracy remains unclear. Recently, DP has shown promise in supporting differential diagnoses^24^, and depressive symptom detection^25–27^. Studies have used *active DP* (e.g., diary entries, open-ended text entries^28–30^, self-report mood log^30,31^, social media behaviours^32–35^) and *passive DP* (e.g., movement patterns, activity levels^36,37^, heart rate^36^, objective sleep measures^38,39^) to support PPD symptoms prediction. However, the predictive performance varies by time frames^29,36^, and modality combination^29,40^, and some modalities (as mobility patterns^41^, step counts^36^, sleep efficiency^38^, content of images posted on social media^32^, heart rate^36^) show no to limited predictive value. This review summarizes available evidence on DP for PPD prediction and identification, including data types, timing, analysis methods, and predictive metrics.

## Methods

This review was conducted following the Preferred Reporting Items for Systematic Reviews and Meta-Analyses (PRISMA) guidelines (supplementary table 1),^42^, and was registered with PROSPERO (CRD42023461325). Six databases, including PubMed, Web of Science, PsycINFO, CINAHL, Cochrane Trials, and Scopus, were searched on the 11^th^ of October 2023 and updated in March 2025. Search results were imported into Rayyan^43^ for duplicate removal and screening. An additional search using Google Scholar was conducted to include more studies before preparing manuscript.

### Search strategy and eligibility criteria

Two keyword groups were used: for the perinatal period (e.g., “antenatal,” “prenatal,” “postpartum,” “perinatal,” “peripartum,” and “maternal depression”) and another one for DP terms (e.g., “digital phenotyping,” “wearable device,” “active digital data,” “passive digital data,” “smartphone,” “mobile application,” “real-time data,” “ecological momentary assessment,” “text message,” “digital behaviours,” “social media,” and “fitness device”). Boolean logic and search syntex are detailed in supplementary table 2. A three-step process (titles, abstracts, full articles) was independently conducted by the first and last authors, retaining articles if either deemed them relevant. Additional studies were identified via Google Scholar (first 30 pages) and reference checks. Inclusion criteria (based on PICO^44^) covered participants during pregnancy and 12 months postpartum, andsStudies providing active and/or passive digital information for PPD prediction or identification. Excluded were protocols, reviews, feasibility studies, and those applying digital applications for interventions or single-time-point questionnaires (supplementary table 3).

### Data extraction

Extracted data included: participant demographics, timeframe, recruitment criteria, data sources (online platforms, health apps, etc.), data types (active or passive data format), PPD assessment tools, analysis techniques, model performance.

### Quality assessment

Quality assessment was evaluated using the Prediction model Risk of Bias Assessment Tool (PROBAST)^45^, a structured tool to assess the risk of bias and the applicability of prediction models. Four domains including participants, independent variables, outcomes, and analyses were assessed independently by two authors, and jointly finalised by three authors. Each domain was rated as low, high, or unclear. Notably, models without external validation were rated high risk unless based on large datasets with internal validation. Studies not reporting prediction results were rated unclear.

## Results

### Study selection

The search process identified 4,356 records from databases including PubMed, Web of Science, PsycINFO, CINAHL, Cochrane Trials, and Scopus. After removing 1,321 duplicates, 3,035 unique records were screened by title, resulting in 109 for abstract screening. The selection process is illustrated in the PRISMA flowchart (Fig. 1).

**Fig. 1.**
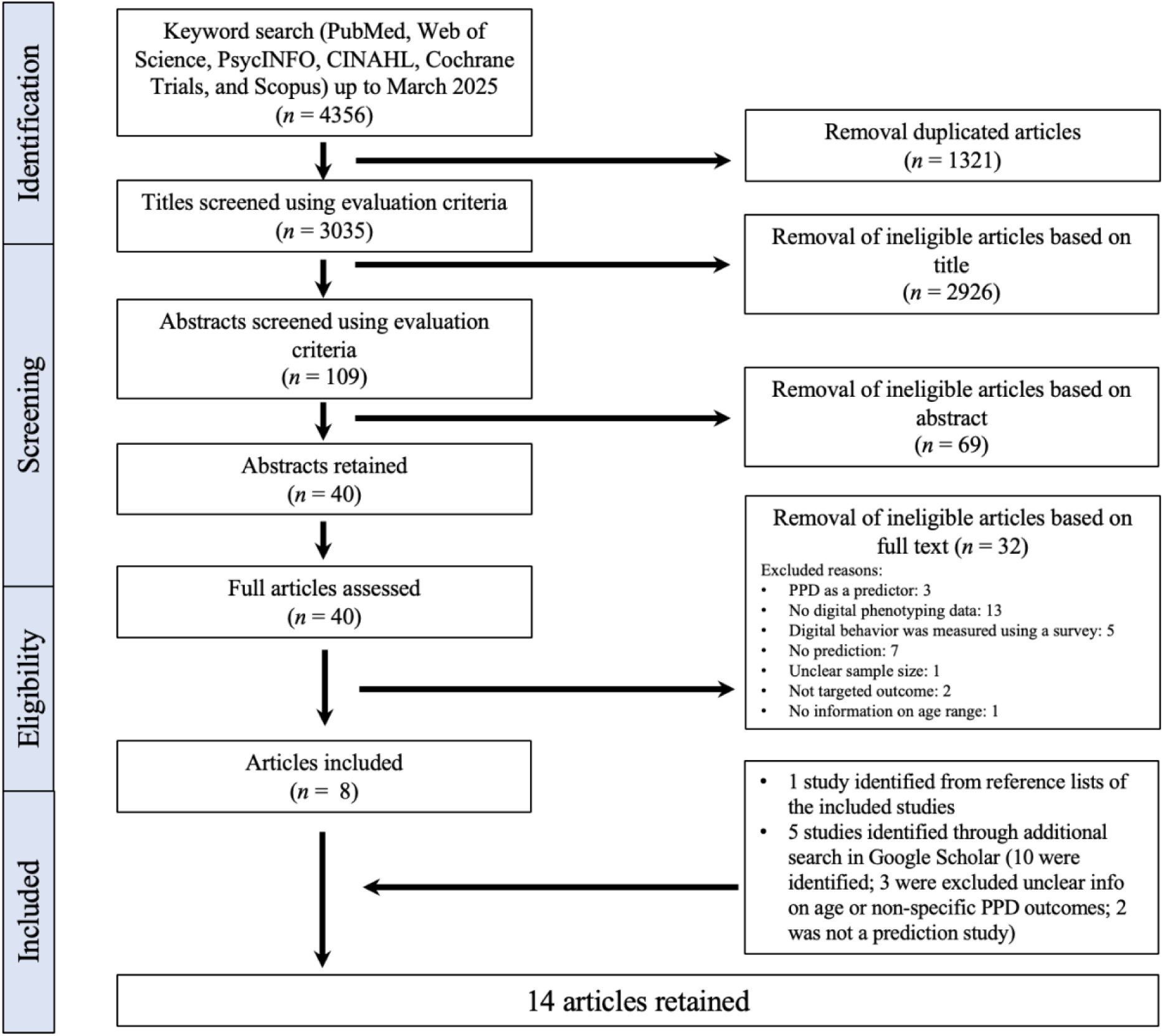
PRISMA flow diagram. This diagram illustrates the study selection process. A total of 4,356 records were identify through searches in seven databases up to March 2025. After screening and applying inclusion criteria, 14 studies were included in the final report.

### Study Characteristics

Table 1 summaries 14 studies conducted from 2014 to 2025 (March) from the USA (k = 8), Sweden (k = 3), Canada (k = 1), China (k = 1), and Germany (k = 1). Most employed observational or cross-sectional designs, with sample sizes from 56 to 2,062 and participants aged 14 to 45. While the majority were Caucasian, some studies included Asian and African populations. Data sources included social media^29,32,33,39^, outpatient clinics^29,37^, university hospitals^30,31,38,40^, mobile applications^41^, perinatal health care centres^29,46^, and a national research dataset^36,47^. Three studies focused on the antenatal period, three on the postnatal period, and eight included both.

**Table 1.**
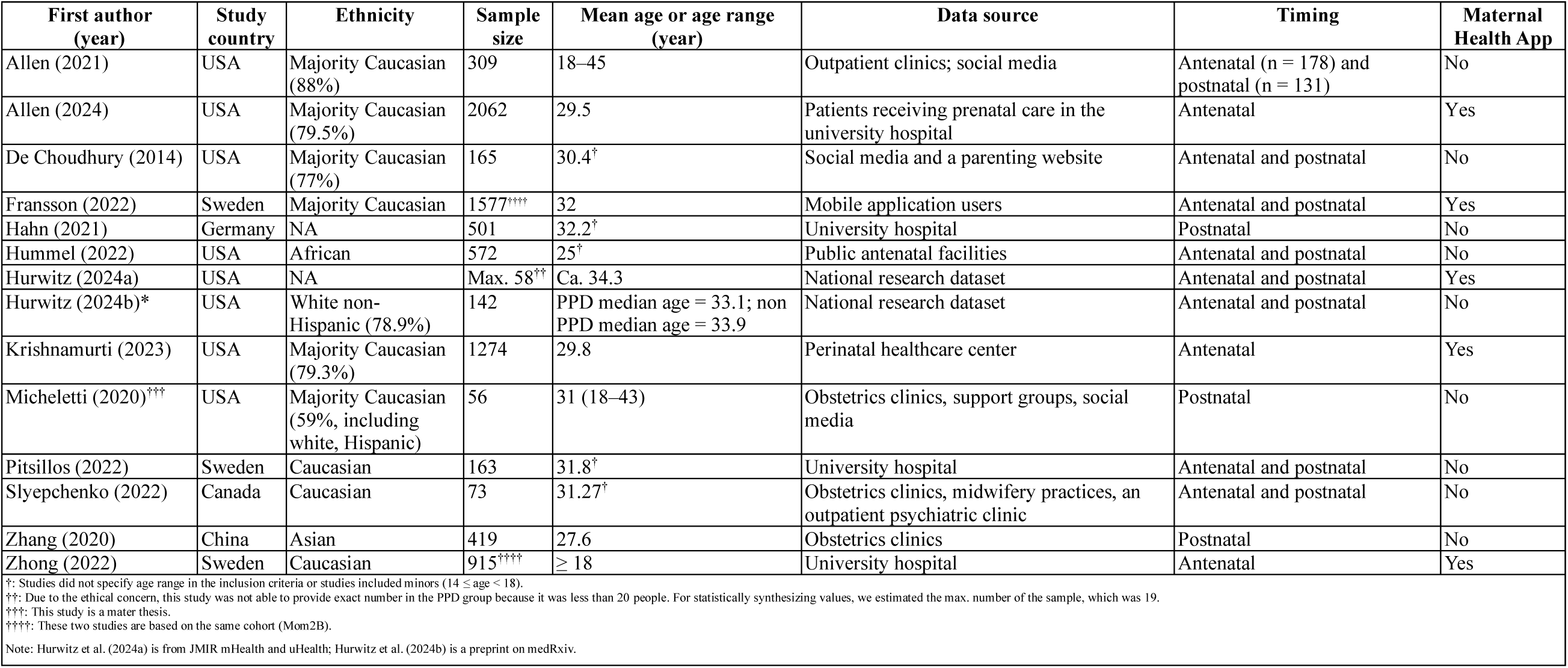
Study characteristics and summary.

### Types of DP data and their predictive results for PPD

Fig. 2 shows key timeframes and measurement types across different studies. Among the 14 studies on PPD, the Edinburgh Postnatal Depression Scale (EPDS) was the primary outcome measure in 9 studies, making it the most commonly used tool. The remaining outcome measures include Patient Health Questionnaire (PHQ-9), Hamilton Depression Rating Scale (HDRS), clinical diagnosis, PHQ-4, clinical interviews, Montgomery-Asberg Depression Rating Scale (MADRS), diagnosis, and Postpartum Depression Predictors Inventory-Revised (PDPI-R). DP modalities were used, including sleep, physical activity, text, social media behaviour, SMS behaviour, ecological momentary assessment, self-report survey, and mood log. Four studies ^29,32,37,46^ primarily utilized DP data for prediction while other studies incorporated additional data sources into the prediction models, such as sociodemographic data^28,31,33,38^, anamnestic data^31^, electronic health records^36^, self-reported symptoms^28,31,40^, ongoing mental health problems^38^, and self-reported social support^39^. While some DP modalities showed predictive values, combining multiple modalities generally improved model performance. Due to variability in parameters and methodologies, results could not be statistically synthesized across studies. Fig. 3 highlights the top-performing modalities or combinations, and Table 2 details methods and results. Key findings are also summarized in supplementary table 4.

**Fig. 2.**
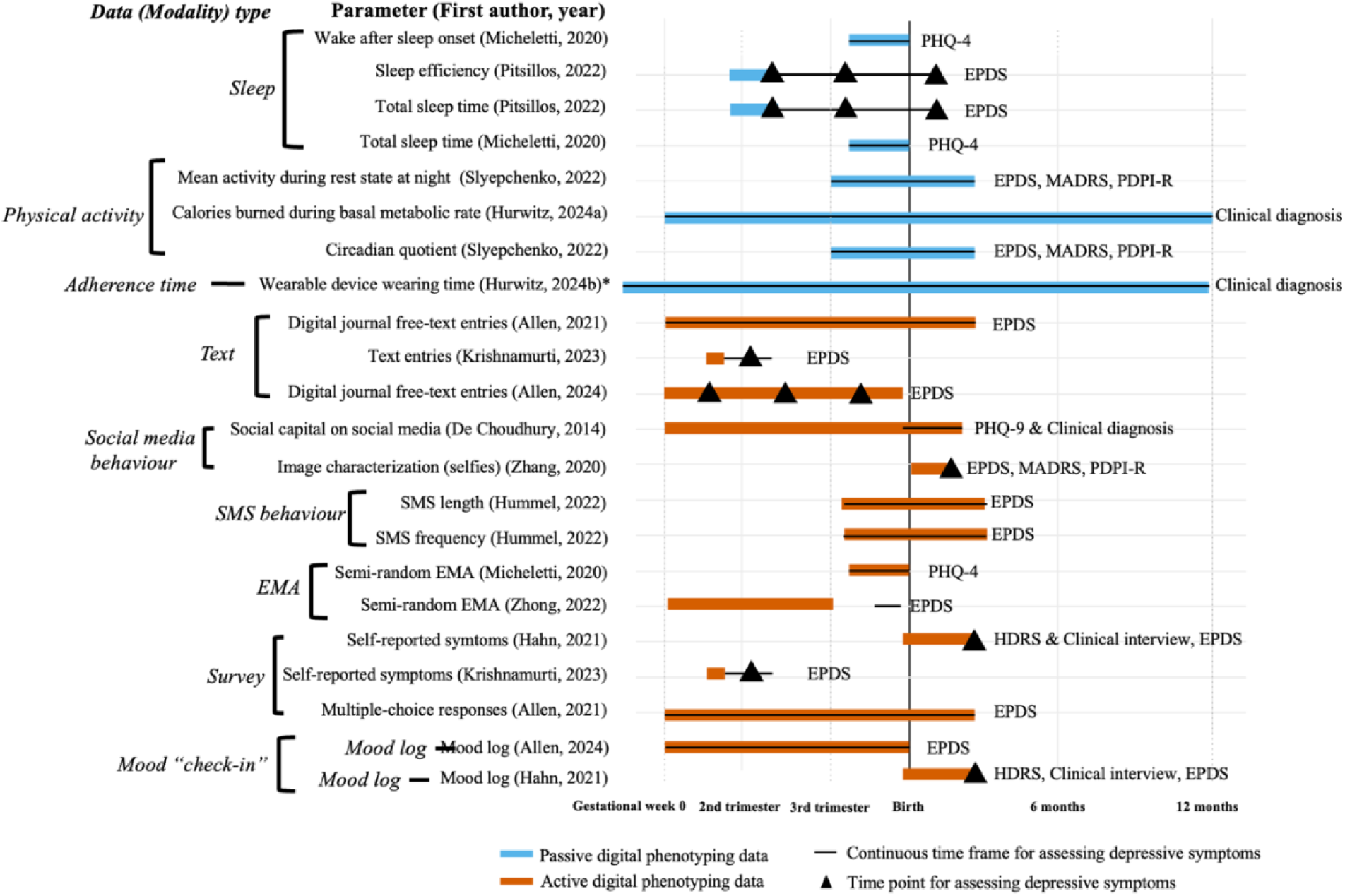
Key timeframes and digital phenotyping data types across studies. Illustration of the included studies, categorized by different digital phenotyping data (modality) types and measurement timing. Abbreviations: EMA (Ecological Momentary Assessment), EPDS (Edinburgh Postnatal Depression Scale), HDRS (Hamilton Depression Rating Scale), MADRS (Montgomery-Asberg Depression Rating Scale), PDPI-R (Postpartum Depression Predictors Inventory-Revised), PHQ (Patient Health Questionnaire), and SMS (Short Message Service). Note: Hurwitz et al. (2024a) is from JMIR mHealth and uHealth; Hurwitz et al. (2024b) is a preprint on medRxiv.

**Fig. 3.**
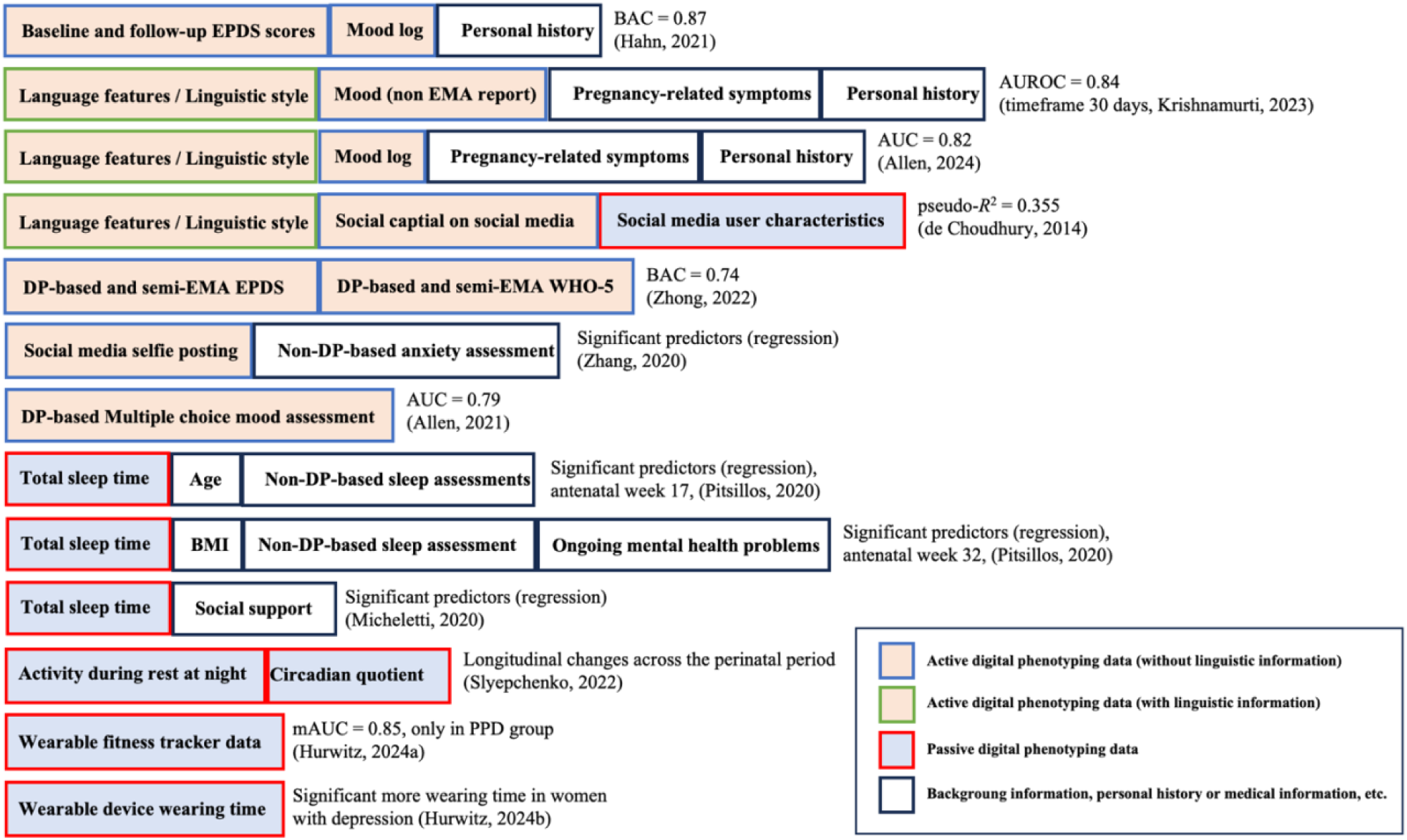
Best modality or combination of modalities across studies. This figure presents the best combinations of data modalities identified across various studies, with each entry represented by the first author’s name and publication year. Red-bordered, light blue boxes indicate passive digital phenotyping data. Green-bordered, light orange boxes denote active digital phenotyping data, further divided into blue (non-language-specific information) and green (language-focused information, such as open-ended text entries and journals, often processed using natural language models). Black-bordered, white boxes represent non-digital phenotyping data, including background information, medical records, personal history, and other conventional clinical data. Abbreviations: AUC (Area under the curve), AUROC (Area under the receiver operating characteristic curve), BAC (Balanced accuracy), BMI (Body mass index), DP (Digital phenotyping), EMA (Ecological momentary assessment), EPDS (Edinburgh Postnatal Depression Scale), mAUC (Mean area under the curve), PPD (Peripartum depression), WHO-5 (World Health Organization 5-Item Well-Being Index). Note: Hurwitz et al. (2024a) is from JMIR mHealth and uHealth; Hurwitz et al. (2024b) is a preprint on medRxiv.

**Table 2.**
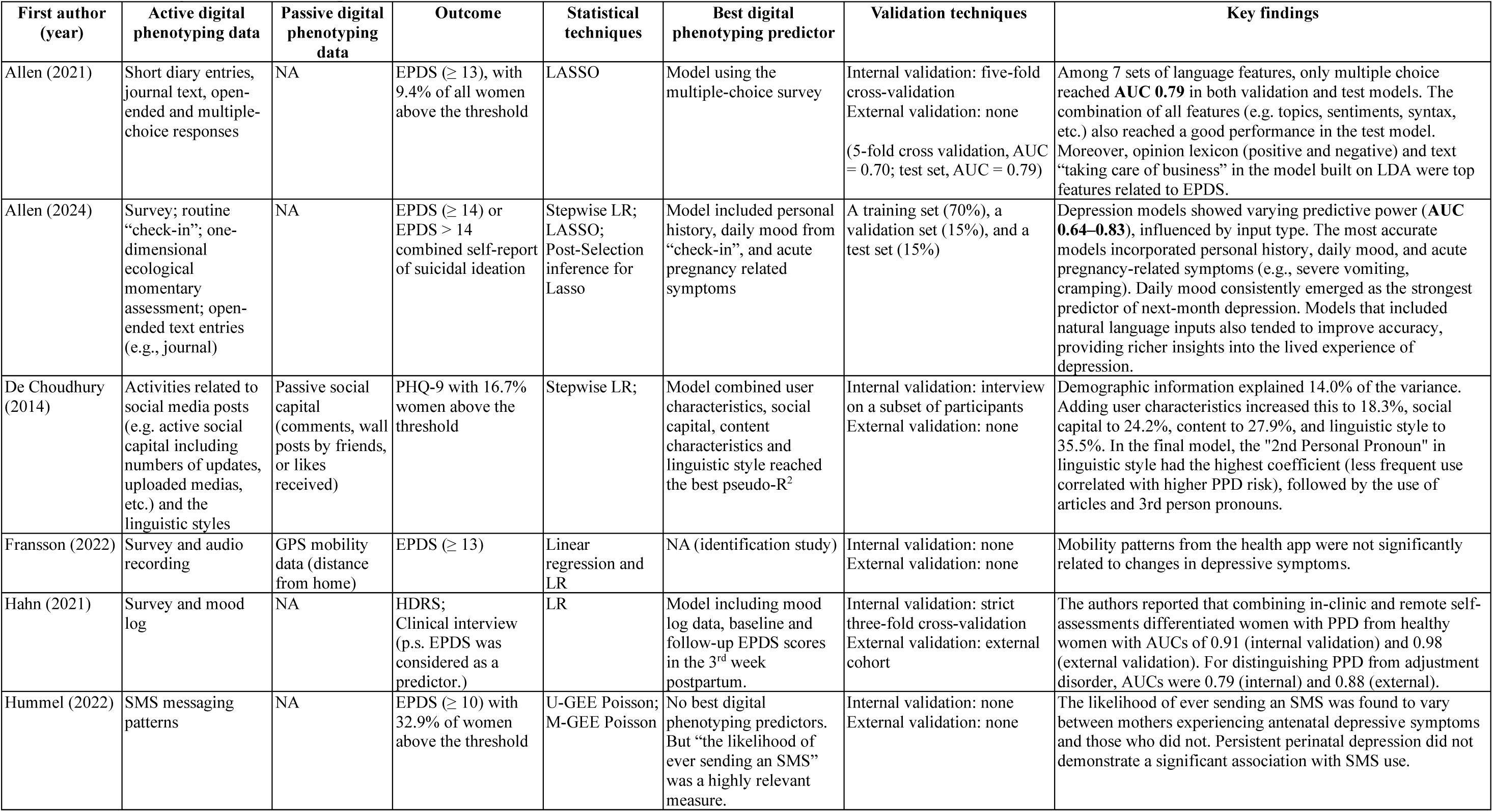
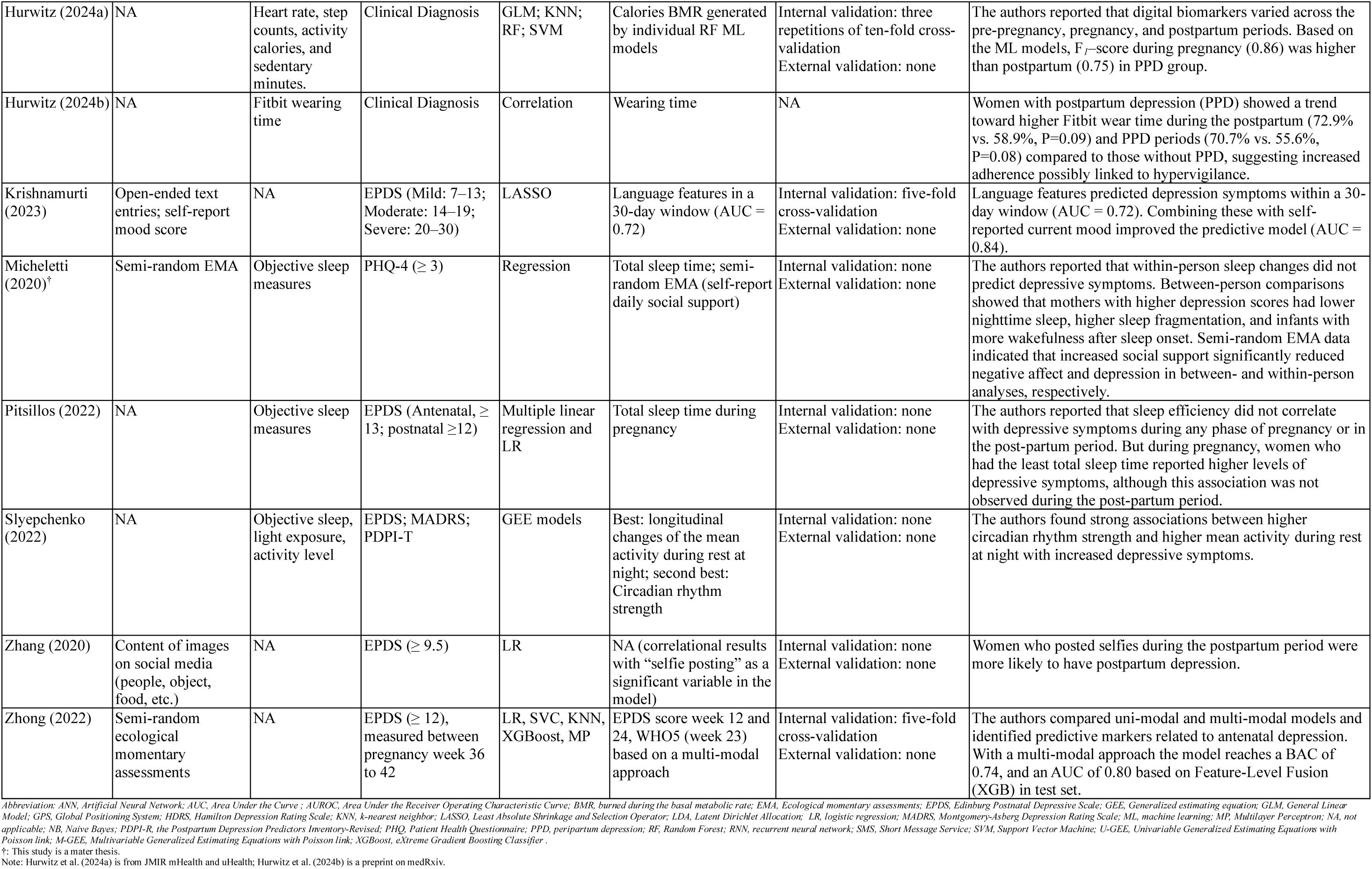
Summary of methodological details and results across 14 studies.

| First author (year) | Active digital phenotyping data | Passive digital phenotyping data | Outcome | Statistical techniques | Best digital phenotyping predictor | Validation techniques | Key findings |
| --- | --- | --- | --- | --- | --- | --- | --- |
| Allen (2021) | Short diary entries, journal text, open-ended and multiple-choice responses | NA | EPDS ($\geq 13$), with 9.4% of all women above the threshold | LASSO | Model using the multiple-choice survey | Internal validation: five-fold cross-validation<br>External validation: none<br>(5-fold cross validation, AUC = 0.70; test set, AUC = 0.79) | Among 7 sets of language features, only multiple choice reached **AUC 0.79** in both validation and test models. The combination of all features (e.g. topics, sentiments, syntax, etc.) also reached a good performance in the test model. Moreover, opinion lexicon (positive and negative) and text “taking care of business” in the model built on LDA were top features related to EPDS. |
| Allen (2024) | Survey; routine “check-in”; one-dimensional ecological momentary assessment; open-ended text entries (e.g., journal) | NA | EPDS ($\geq 14$) or EPDS > 14 combined self-report of suicidal ideation | Stepwise LR; LASSO; Post-Selection inference for Lasso | Model included personal history, daily mood from “check-in”, and acute pregnancy related symptoms | A training set (70%), a validation set (15%), and a test set (15%) | Depression models showed varying predictive power (**AUC 0.64–0.83**), influenced by input type. The most accurate models incorporated personal history, daily mood, and acute pregnancy-related symptoms (e.g., severe vomiting, cramping). Daily mood consistently emerged as the strongest predictor of next-month depression. Models that included natural language inputs also tended to improve accuracy, providing richer insights into the lived experience of depression. |
| De Choudhury (2014) | Activities related to social media posts (e.g. active social capital including numbers of updates, uploaded medias, etc.) and the linguistic styles | Passive social capital (comments, wall posts by friends, or likes received) | PHQ-9 with 16.7% women above the threshold | Stepwise LR; | Model combined user characteristics, social capital, content characteristics and linguistic style reached the best pseudo-$R^2$ | Internal validation: interview on a subset of participants<br>External validation: none | Demographic information explained 14.0% of the variance. Adding user characteristics increased this to 18.3%, social capital to 24.2%, content to 27.9%, and linguistic style to 35.5%. In the final model, the “2nd Personal Pronoun” in linguistic style had the highest coefficient (less frequent use correlated with higher PPD risk), followed by the use of articles and 3rd person pronouns. |
| Fransson (2022) | Survey and audio recording | GPS mobility data (distance from home) | EPDS ($\geq 13$) | Linear regression and LR | NA (identification study) | Internal validation: none<br>External validation: none | Mobility patterns from the health app were not significantly related to changes in depressive symptoms. |
| Hahn (2021) | Survey and mood log | NA | HDRS; Clinical interview (p.s. EPDS was considered as a predictor.) | LR | Model including mood log data, baseline and follow-up EPDS scores in the 3<sup>rd</sup> week postpartum. | Internal validation: strict three-fold cross-validation<br>External validation: external cohort | The authors reported that combining in-clinic and remote self-assessments differentiated women with PPD from healthy women with AUCs of 0.91 (internal validation) and 0.98 (external validation). For distinguishing PPD from adjustment disorder, AUCs were 0.79 (internal) and 0.88 (external). |
| Hummel (2022) | SMS messaging patterns | NA | EPDS ($\geq 10$) with 32.9% of women above the threshold | U-GEE Poisson; M-GEE Poisson | No best digital phenotyping predictors. But “the likelihood of ever sending an SMS” was a highly relevant measure. | Internal validation: none<br>External validation: none | The likelihood of ever sending an SMS was found to vary between mothers experiencing antenatal depressive symptoms and those who did not. Persistent perinatal depression did not demonstrate a significant association with SMS use. |
| Hurwitz (2024a) | NA | Heart rate, step counts, activity calories, and sedentary minutes. | Clinical Diagnosis | GLM; KNN; RF; SVM | Calories BMR generated by individual RF ML models | Internal validation: three repetitions of ten-fold cross-validation<br>External validation: none | The authors reported that digital biomarkers varied across the pre-pregnancy, pregnancy, and postpartum periods. Based on the ML models, $F_1$-score during pregnancy (0.86) was higher than postpartum (0.75) in PPD group. |
| Hurwitz (2024b) | NA | Fitbit wearing time | Clinical Diagnosis | Correlation | Wearing time | NA | Women with postpartum depression (PPD) showed a trend toward higher Fitbit wear time during the postpartum (72.9% vs. 58.9%, $P=0.09$) and PPD periods (70.7% vs. 55.6%, $P=0.08$) compared to those without PPD, suggesting increased adherence possibly linked to hypervigilance. |
| Krishnamurti (2023) | Open-ended text entries; self-report mood score | NA | EPDS (Mild: 7–13; Moderate: 14–19; Severe: 20–30) | LASSO | Language features in a 30-day window (AUC = 0.72) | Internal validation: five-fold cross-validation<br>External validation: none | Language features predicted depression symptoms within a 30-day window (AUC = 0.72). Combining these with self-reported current mood improved the predictive model (AUC = 0.84). |
| Micheletti (2020)<sup>†</sup> | Semi-random EMA | Objective sleep measures | PHQ-4 ($\geq 3$) | Regression | Total sleep time; semi-random EMA (self-report daily social support) | Internal validation: none<br>External validation: none | The authors reported that within-person sleep changes did not predict depressive symptoms. Between-person comparisons showed that mothers with higher depression scores had lower nighttime sleep, higher sleep fragmentation, and infants with more wakefulness after sleep onset. Semi-random EMA data indicated that increased social support significantly reduced negative affect and depression in between- and within-person analyses, respectively. |
| Pitsillos (2022) | NA | Objective sleep measures | EPDS (Antenatal, $\geq 13$; postnatal $\geq 12$) | Multiple linear regression and LR | Total sleep time during pregnancy | Internal validation: none<br>External validation: none | The authors reported that sleep efficiency did not correlate with depressive symptoms during any phase of pregnancy or in the post-partum period. But during pregnancy, women who had the least total sleep time reported higher levels of depressive symptoms, although this association was not observed during the post-partum period. |
| Slyepchenko (2022) | NA | Objective sleep, light exposure, activity level | EPDS; MADRS; PDPI-T | GEE models | Best: longitudinal changes of the mean activity during rest at night; second best: Circadian rhythm strength | Internal validation: none<br>External validation: none | The authors found strong associations between higher circadian rhythm strength and higher mean activity during rest at night with increased depressive symptoms. |
| Zhang (2020) | Content of images on social media (people, object, food, etc.) | NA | EPDS ($\geq 9.5$) | LR | NA (correlational results with “selfie posting” as a significant variable in the model) | Internal validation: none<br>External validation: none | Women who posted selfies during the postpartum period were more likely to have postpartum depression. |
| Zhong (2022) | Semi-random ecological momentary assessments | NA | EPDS ($\geq 12$), measured between pregnancy week 36 to 42 | LR, SVC, KNN, XGBoost, MP | EPDS score week 12 and 24, WHO5 (week 23) based on a multi-modal approach | Internal validation: five-fold cross-validation<br>External validation: none | The authors compared uni-modal and multi-modal models and identified predictive markers related to antenatal depression. With a multi-modal approach the model reaches a BAC of 0.74, and an AUC of 0.80 based on Feature-Level Fusion (XGB) in test set. |

Abbreviation: ANN, Artificial Neural Network; AUC, Area Under the Curve; AUROC, Area Under the Receiver Operating Characteristic Curve; BMR, burned during the basal metabolic rate; EMA, Ecological momentary assessments; EPDS, Edinburg Postnatal Depressive Scale; GEE, Generalized estimating equation; GLM, General Linear Model; GPS, Global Positioning System; HDRS, Hamilton Depression Rating Scale; KNN, k-nearest neighbor; LASSO, Least Absolute Shrinkage and Selection Operator; LDA, Latent Dirichlet Allocation; LR, logistic regression; MADRS, Montgomery-Asberg Depression Rating Scale; ML, machine learning; MP, Multilayer Perceptron; NA, not
applicable; NB, Naive Bayes; PDPI-R, the Postpartum Depression Predictors Inventory-Revised; PHQ, Patient Health Questionnaire; PPD, peripartum depression; RF, Random Forest; RNN, recurrent neural network; SMS, Short Message Service; SVM, Support Vector Machine; U-GEE, Univariable Generalized Estimating Equations with Poisson link; M-GEE, Multivariable Generalized Estimating Equations with Poisson link; XGBoost, eXtreme Gradient Boosting Classifier .
†: This study is a master thesis.
Note: Hurwitz et al. (2024a) is from JMIR mHealth and uHealth; Hurwitz et al. (2024b) is a preprint on medRxiv.

### Findings from passive DP data

The included studies reveal four major subgroups of passive digital data (Fig. 2). The first focuses on objective sleep measurements,^38,39^ highlighting key parameters like sleep patterns and total sleep time. Micheletti et al. monitored sleep continuously over 72 hours during the third trimester (31-41 weeks) and found that within-person sleep changes did not predict depressive symptoms assessed six times during that week. However, between-person comparisons indicated that lower nighttime sleep and higher sleep fragmentation were linked to higher depression scores in the third trimester.^39^ Similarly, Pitsillos et al.^38^ reported that shorter total sleep time between 11–19 gestational weeks was significantly associated with increased depressive symptoms at 16 and 32 weeks, though not at six weeks postpartum^38^. The second subgroup involved physical activity data, including movement patterns (Global Positioning System, GPS)^41^, heart rate, step counts, activity calories, sedentary minutes^36^, and night activity level^37^. Hurwitz et al.^36^ analysed Fitbit data from two years pre-pregnancy to two years postpartum and found significant associations between calories burned at resting state and clinical PPD diagnoses at two years postpartum. However, other metrics such as step count, active minutes, calories burned during physical activity, total distance and activity type, did not predict PPD^36^. In another study, Slyepchenko et al.^37^collected physical activity and sleep data at three time points, late pregnancy, 1-3 weeks postpartum, and 6-12 weeks postpartum, and found that nighttime activity and disrupted daily rhythms were robust predictors of PPD symptoms^37^. In contrast, GPS data collected from early pregnancy through one year postpartum showed no significant association with PPD outcomes^41^. The third subgroup, passive social capital, included metrics such as comments, likes, and wall posts on social media^33^. De Choudhury et al.^33^ reported that the combination of demographic info, user characteristics and social capital measures predict PPD the best, with social capital metrics (numbers of updates, comments, etc.) contributing substantial predictive value. However, these metrics were not analyzed in isolation but rather as part of a broader model incorporating active DP data, and are therefore categorized under “active DP” in Fig. 2 and Fig. 3. Adherence time, the fourth subgroup capturing wearable device wear duration, showed that women with postpartum depression (PPD) tended to wear their devices more consistently during the postpartum and PPD periods. This pattern may reflect behavioral markers such as hypervigilance, which could support early detection and monitoring of mood disorders^47^.

### Findings from active DP data

Six subgroups (Fig. 2) of active DP data were identified: text entries,^28,29^ social media behaviours^32,33^, semi-random Ecological Momentary Assessments (EMAs)^39,40^, short message service behaviour^46^, survey (self-reported)^28,29,31^, and mood log^30,31^. Text entries were used in two studies. Allen et al. ^29^ collected text entries throughout pregnancy and up to 12 weeks postpartum, finding that negative sentiment, physical exhaustion, and lack of positive affect in text were significantly associated with depressive symptoms. The predictive model improved when survey data were included^29^. Similarly, Krishnamurti et al. analyzed text entries and survey data from the first trimester found that language features, such as sentiment fluctuation, mental health-related language (e.g., “depressed”, “suicidal”, “trauma”), pregnancy-related topics, and the use of first-person plural pronouns (e.g., “we”, “our”, “us”), predicting PPD within a 30-day window. Predictive accuracy increased when combined with self-reported mood data^28^. Social media behavior were examined in two studies. De Choudhury et al.^33^ monitored social media behavior from early pregnancy to 10 weeks postpartum and found that increased social isolation and reduced social capital were strong predictors of PPD symptoms^33^. Zhang et al.^32^ observed that mothers who posted more selfies of themselves or their children were more likely to experience depressive symptoms at six weeks postpartum, while other image types (e.g., memes) were not predictive^32^. Short message service (SMS) behavior was assessed by Hummel et al.^46^ who found that fewer and shorter messages during late pregnancy and at 14 weeks postpartum were associated with depressive symptoms. However, persistent depression did not significantly affect the likelihood of sending messages^46^. Two studies used survey data integrated using semi-random EMA for prediction^39,40^. Micheletti et al.^39^ conducted semi-random EMAs six times daily over seven days in late pregnancy, finding that increased social support significantly reduced negative affect and depression in both between- and within-person analyses^39^. Zhong et al.^40^ collected semi-random EMA-based survey data during the first and second trimesters and found that EPDS scores, WHO-5, stress, and anxiety levels were significant predictors of PPD in the third trimester^40^. Self-report surveys were often integrated with other data types to enhance prediction. For example, Hahn et al.^31^ utilized mood logs recorded twice daily from birth to 12 weeks postpartum and found significant associations with PPD outcomes. Combining mood logs with clinical assessments (HDRS, EPDS, and interviews) improved model accuracy and effectively distinguished between women with and without depressive symptoms^31^.

### Statistical approaches

The statistical methods employed across the studies are summarized in Fig. 4. For well-structured datasets with predefined assumptions, various statistical methods can be employed to model relationships between dependent and independent variables and assess statistical significance. These methods include Linear Regression, Multiple Linear Regression (MLR), the General Linear Model (GLM), and Generalized Estimating Equations (GEE). Furthermore, Poisson Regression Models were used for count data, with Univariable and Multivariable Generalized Estimating Equations (U-GEE and M-GEE) applied to assess single and multiple predictors, respectively. The Machine Learning (ML) approaches encompass a diverse range of algorithms developed to learn patterns from data and make predictions. They can be further divided into algorithms designed for outcome prediction and those focused on regularization and feature selection. The ML for outcome prediction category includes algorithms like Logistic Regression (LR), Support Vector Machines (SVM), k-Nearest Neighbor (KNN), Decision Trees (DT), including Random Forest (RF) and eXtreme Gradient Boosting Classifier (XGBoost), as well as Artificial Neural Networks (ANNs), including Recurrent Neural Networks (RNN), and simple architectures such as Multilayer Perceptron (MP). The ML for regularization and feature selection category is represented by the Least Absolute Shrinkage and Selection Operator (LASSO), which is particularly useful for high-dimensional data^48^ where the number of features may be large relative to the number of observations. Table 2 summarizes included studies and their statistical approaches.

**Fig. 4.**
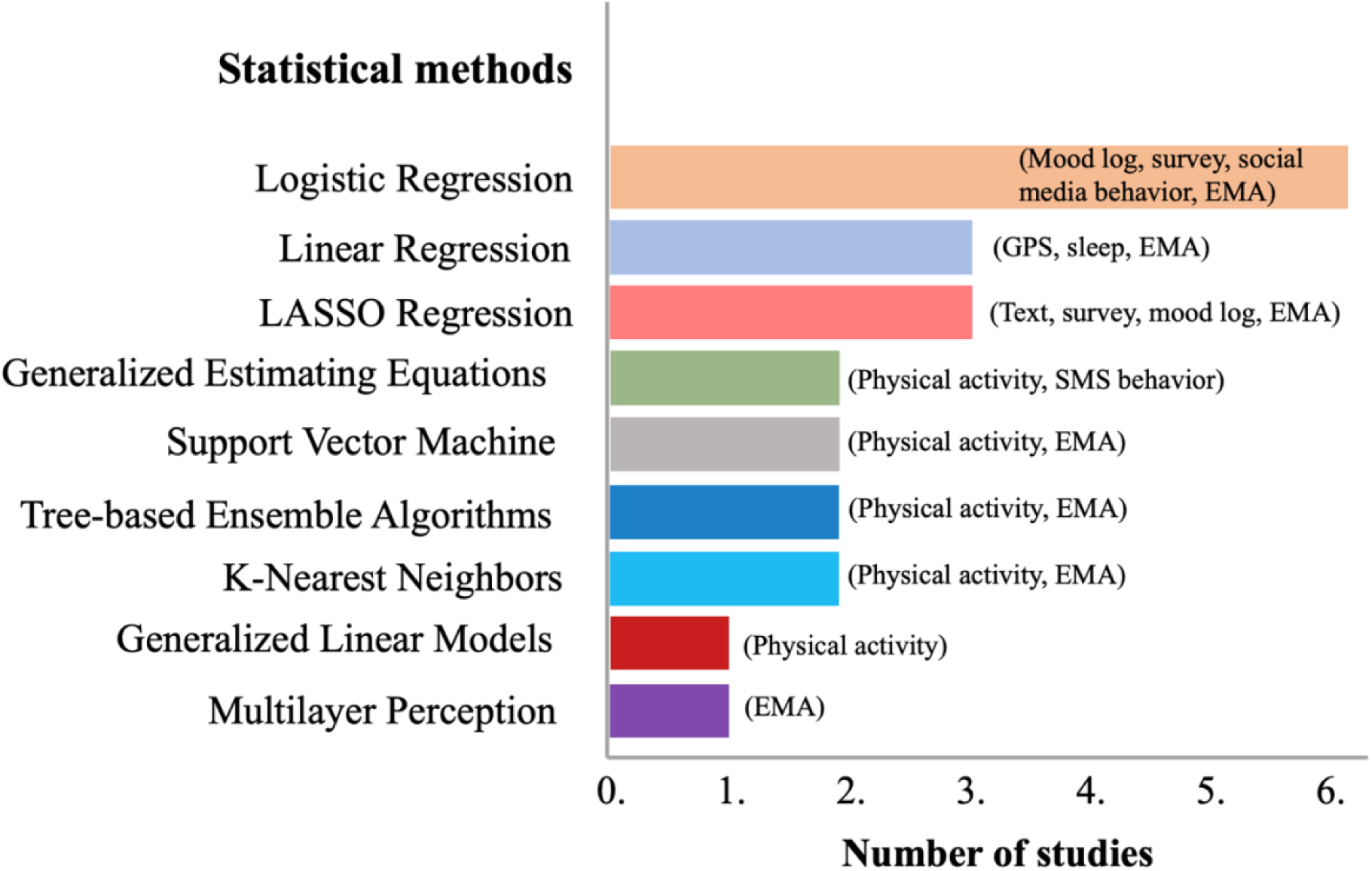
Key statistical methods across studies. Illustration of statistical methods and modalities used in the included studies. The figure presents an overview of the statistical methods applied across the studies. The X-axis indicates the number of studies employing each statistical method, while the Y-axis lists the various methods used. The text content within each bar denotes the different digital phenotyping modalities utilized, not their frequency of use. Logistic regression is the most frequently applied method. Abbreviations include LASSO (Least Absolute Shrinkage and Selection Operator), GEE (Generalized Estimating Equations), and XGBoost (eXtreme Gradient Boosting Classifier). Modalities: GPS, Global Positioning System; EMA, Ecological Momentary Assessment; SMS, Short Message Service. For detailed information, please refer to Table 2.

Linear regression was applied to analyze independent variables such as GPS data^41^ and sleep data^38,39^. The performance of these models varied significantly based on the inclusion of background information and the specific variables utilized in the analysis. GPS-based distance from home showed no significant link to depressive symptoms, indicating limited predictive value^41^. In contrast, one study found significant associations between total sleep time, wake after sleep onset and sleep fragmentation during pregnancy and depressive symptoms^39^. Some additional factors contributing to this model were infant sleep hours, and social support^39^. This study used Multiple Linear Regression and found a significant correlation between total sleep time during pregnancy and depressive symptoms, however no significant relationship was found with sleep efficiency^38^. Furthermore, Poisson Regression Models revealed that fewer and shorter SMS messages were sent by women with antenatal and persistent perinatal depressive symptoms compared to healthy women. GEE models demonstrated that higher circadian rhythm strength and increased nighttime activity were significant predictors of depressive symptoms during the perinatal period^37^.

Machine learning approaches were used in several studies to predict depressive symptoms, employing various algorithms for data analysis. Logistic Regression was the most common technique (k = 6), effectively analyzing data types such as social media behaviour^32,33^, mood logs^31^, mobility patterns^41^, and semi-random EMA surveys^40^. For example, stepwise logistic regression accounted for 35.5% of the variance in depressive symptoms by incorporating demographics, user characteristics, content features, social capital, and linguistic styles^33^. Another study found a significant link between posting selfies and peripartum depression, explaining only 20% of the variance^32^. Logistic regression also demonstrated high predictive accuracy in mood logs and surveys, with AUC values of 0.91 and 0.98.^31^ The Least Absolute Shrinkage and Selection Operator (LASSO) Regression, used in two studies to analyze language features from text entries, achieved AUCs of 0.72, improving to 0.84 with self-reported mood data^28^, and 0.79 with multiple-choice surveys combined with free-text responses^29^. Random Forest models were applied in one study to analyze energy expenditure data, achieving AUCs of 0.86 during pregnancy and 0.75 postpartum^36^. However, logistic regression found no significant association between GPS-based distance from home and depressive symptoms^41^.

Multimodal approaches, utilizing various methods like LR, SVM, KNN, MP, and XGBoost, achieved an AUC of 0.83 by integrating diverse longitudinal data^40^. enhancing predictive accuracy. Among unimodal models, XGBoost performed best with an AUC of 0.81^40^. Overall, machine learning techniques exhibited varying efficacy; the best models depended on data type and structure. Logistic Regression excelled with structured data like mood logs and surveys, while LASSO Regression was more effective for high-dimensional, unstructured data. Additionally, multimodal approaches and Random Forest models for analyzing physiological data demonstrated significant effectiveness.

Among the included studies, only one addressed missing data handling^40^. Regarding model validation, only one out of seven studies conducted both internal and external validation. This study, which analyzed mood logs using Logistic Regression^31^, reported high predictive accuracy, enhancing its findings’ robustness. In contrast, five studies relied solely on internal validation, analyzing social media behaviour^33^, image characterization with Logistic Regression^32^, language features from text entries with LASSO Regression^28,29^, and physical activity data, such as digital biomarkers with Random Forest models^36^.

### Risk of bias and applicability assessment (PROBAST)

The details of PROBAST analysis including 11 studies are listed in supplementary table 5 and the results are summarized in Fig. 5. Three studies were excluded from the prediction quality assessment as they did not meet the design criteria required for PROBAST evaluation. Most of the studies included in the systematic review were classified as having a high risk of bias overall. Ten studies were rated as high risk in the analysis domain, primarily due to inadequate information on missing data handling and the absence of validation procedures. Additionally, in the participant domain, one study had a group with fewer than 20 participants, and another study failed to report the inclusion and exclusion criteria. For overall applicability, most of the studies exhibited low concern. However, one study was assessed as “unclear” due to the independent variable, and one study raised concern due to its very small sample size.

**Fig. 5.**
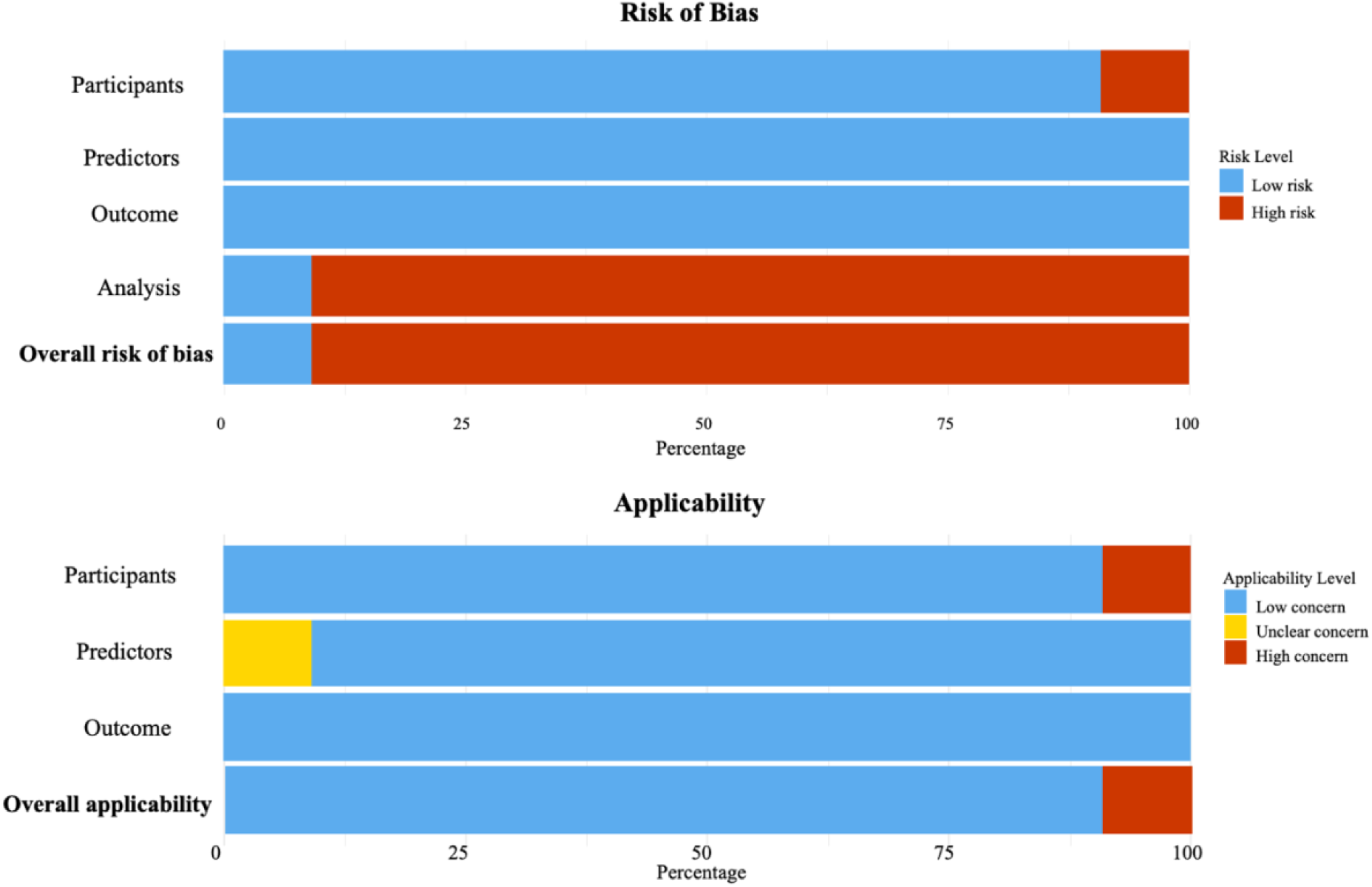
Risk of bias and applicability assessment based on the Prediction model Risk Of Bias ASsessment Tool (PROBAST). Results reflect the 11 studies that reported predictive modeling. Studies reporting only associations were excluded, as PROBAST is not applicable to those designs.

## Discussion

This review is the first to identify key DP variables relevant to the early identification and prediction of PPD, summarizing evidence from 14 studies. While one-third of the studies relied mainly on DP data, most incorporated additional information, such as sociodemographic data, self-reported symptoms, and other relevant factors, into their prediction models. Furthermore, studies reported moderate to high predictive performance often leverage the background and symptom-related information. Most remain in the model development phase, contributing to potential heterogeneity. While several modalities show promise in predicting depressive symptoms, the current literature still reveals a significant gap in translating these findings into large-scale clinical applications. This gap is often driven by challenges in data generalizability, the need for robust internal and external validation, and the complexities of integrating diverse data sources into routine clinical practice. Following this overview, we will discuss some important aspects critical to advancing this field.

### Opportunities for addressing missing data in DP

Despite the promise of DP for predicting depressive symptoms, it’s crucial to acknowledge the risks and biases in data collection and analysis. The included studies exhibited a high risk of bias, particularly in handling missing data and model validation. While DP methods can generate extensive data, concerns about data quality and missing values can lead to inaccurate measurements and misleading results—issues often overlooked in these studies. For instance, a recent study found that after just three days of inactivity, passive DP data coverage dropped by an average of 19%. Study durations ranged from 4 to 52 weeks, and lower data coverage correlated with less accurate results^49^. Missing values can stem from various factors, including low acceptance of digital collection, device malfunctions, connectivity issues, user disengagement, participant fatigue, and unaddressed software updates. Studies that collect DP data from multiple sources (e.g., GPS, accelerometers, surveys) may encounter challenges in integrating this data, which can result in loss or inconsistent recording of information from some modalities. Identifying these types of missingness is essential, as each may require different statistical approaches^50^.

### Enhancing model validation and actively dealing with data heterogeneity

Another issue contributing to a high risk of bias is the lack of model validation. Among the studies reviewed, only Hahn et al.^31^ conducted both internal and external validations. The lack of model validation in both regression and machine learning methods raise critical concerns about the applicability and robustness of predictive models in healthcare. Validation plays a crucial role in regression analysis and machine learning by ensuring the accuracy and reliability of models. In regression analysis, validation helps uncover biases and inaccuracies in estimating relationships between variables, thereby supporting more reliable clinical interpretations and recommendations. Proper validation ensures that models accurately capture real-world variability, leading to predictions that better reflect patient outcomes across different populations and settings.

### Improving consistency and transparency in reporting DP analyses

Although some studies explore similar aspects of DP, there is a lack of consistent terminology, standardized measures, and analytical procedures. For instance, while analyzing active text entries, studies use natural language processing (NLP) or sentiment analysis, but the methods and tools vary significantly, leading to inconsistent results and difficulties in comparison. Emerging evidence suggests that large language models (LLMs) in NLP can predict and monitor adverse mental health outcomes^51,52^. However, the effectiveness of large language models (LLMs) is highly sensitive to hyperparameter tuning, which is crucial for optimizing model performance and ensuring accurate and reliable predictions. Unfortunately, the included studies did not report detailed information on hyperparameters. Properly adjusting these settings is essential to reduce variability and improve the robustness of findings in DP research. Future research should continue to explore innovative statistical approaches, while maintaining a focus on interpretability, clinical applicability, and generalizability.

### Enhancing interdisciplinary collaboration

Our review showed that integrating background information, medical history, clinical data, personal history, and real-time, continuous symptom monitoring can improve predictive accuracy. While a comprehensive approach can help capture a richer, more personalized understanding of PPD, realizing this potential requires overcoming several challenges. First, data privacy remains a critical concern, as sensitive personal information must be protected without compromising predictive power. Second, interdisciplinary collaboration is vital, for instance, clinicians, data scientists, engineers, and ethicists must work together to create ethically sound, technically robust, and clinically meaningful DP tools.

### Practical implications and future research

Current evidence is insufficient to identify which DP variables are most predictive of peripartum depressive symptoms for clinical use or to determine optimal measurement timing and duration. While key psychosocial risk factors for PPD have been identified, our review suggests that integrating validated assessment tools with semi-random EMA could enhance the ability to capture real-time symptom development, and perhaps identify patterns related to worsening mood. Therefore, conducting a baseline assessment of major psychosocial factors along with passive data collection (e.g., sleep, physical activity) would be valuable. Additionally, ongoing inquiry about emerging risk factors, such as bleeding volume and breastfeeding challenges, is crucial during the peripartum period to ensure comprehensive risk management. These factors often cannot be assessed until after delivery, making it important to maintain open lines of communication with mothers to ensure comprehensive risk management. However, these strategies require robust digital infrastructure and ethical considerations to assist at-risk women effectively. Additionally, caution is needed in integrating DP into clinical practice to avoid exacerbating societal inequalities related to race, gender, and nationality^53^. Future research should focus on the development of a unified framework across disciplines to standardize terminology and methods, incorporating tools like the Clinical Utility Index (CUI) to enhance decision-making. The integration of insights from fields such as psychology, data science, and clinical medicine will facilitate a more cohesive approach to research and allow for the standardization of practices. This unified framework should be accompanied by comprehensive guidelines for reporting results and implementing open science practices, thereby fostering collaboration and ensuring that findings are both comparable and generalizable across different studies and populations.

## Conclusions

In conclusion, digital healthcare applications, particularly those utilizing a multimodal approach, show potential for supporting prediction for PPD. By integrating medical records, background information, and active DP data such as mood logs or semi-random ecological momentary data, these tools can provide a more comprehensive understanding of mental health. For passive data, so far, sleeping seems to be promising. While challenges related to data variability, methodological differences, and bias remain, further research aimed at addressing these issues will be crucial for optimizing the role of digital phenotyping in mental health management.

## Supporting information

Supplementary Table 2. Search boolean

Supplementary Table 3. PICO inclusion and exclusion criteria

Supplementary Table 4. ML matrix reported

## Author contributions

B.Z.K. contributed to data curation, article screening, drafting the first version of the manuscript, and preparation of figures and tables. S.S. assisted with data curation, manuscript editing, and preparation of figures and tables. F.C.P. provided critical feedback. A.B. and A.S. contributed to the conceptualization of the study, provided critical feedback, and supported the project through funding. H.-F.T. conceptualized the study, conducted data curation, participated in article screening, manuscript writing, provided critical feedback, supported the project through funding, and offered supervision and guidance. All authors reviewed and approved the final version of the manuscript.

## Data Availability

This study is a review. All relevant information is available in the Supplementary Material. No participant-level data are applicable or required for availability.

## Acknowledgements

This review project was supported by the STePPS CRE project led by the Parent-Infant Research Institute (PIRI) in Melbourne, Australia. We would like to show our great gratitude to Prof. Jeannette Milgrom, who leads the STePPS CRE project, and Dr. Yafit Hirshler for administrative support.

## References

1 Bai, Y. et al. Prevalence of postpartum depression based on diagnostic interviews: a systematic review and meta-analysis. Depression and Anxiety 2023, 8403222 (2023). 10.1155/2023/8403222

2 Mitchell, A. R. et al. Prevalence of Perinatal Depression in Low-and Middle-Income Countries: A Systematic Review and Meta-analysis. JAMA psychiatry (2023). 10.1001/jamapsychiatry.2023.0069

3 Slomian, J., Honvo, G., Emonts, P., Reginster, J.-Y. & Bruyère, O. Consequences of maternal postpartum depression: A systematic review of maternal and infant outcomes. Women’s Health 15, 1745506519844044 (2019). 10.1177/1745506519844044

4 Rogers, A. et al. Association between maternal perinatal depression and anxiety and child and adolescent development: a meta-analysis. JAMA pediatrics 174, 1082–1092 (2020). 10.1001/jamapediatrics.2020.2910

5 Torous, J., Kiang, M. V., Lorme, J. & Onnela, J.-P. New tools for new research in psychiatry: a scalable and customizable platform to empower data driven smartphone research. JMIR mental health 3, e5165 (2016). 10.2196/mental.5165

6 Benoit, J., Onyeaka, H., Keshavan, M. & Torous, J. Systematic review of digital phenotyping and machine learning in psychosis spectrum illnesses. Harvard Review of Psychiatry 28, 296–304 (2020). 10.1097/HRP.0000000000000268

7 Bloch, M., Daly, R. C. & Rubinow, D. R. Endocrine factors in the etiology of postpartum depression. Comprehensive Psychiatry 44, 234–246 (2003). 10.1016/S0010-440X(03)00034-8

8 O’Hara, M. W. & McCabe, J. E. Postpartum depression: current status and future directions. Annual Review of Clinical Psychology 9, 379–407 (2013). 10.1146/annurev-clinpsy-050212-185612

9 Howard, L. M. et al. Non-psychotic mental disorders in the perinatal period. The Lancet 384, 1775–1788 (2014). 10.1016/S0140-6736(14)61276-9

10 O’Hara, M. W. & Wisner, K. L. Perinatal mental illness: definition, description and aetiology. Best Practice & Research Clinical Obstetrics & Gynaecology 28, 3–12 (2014). 10.1016/j.bpobgyn.2013.09.002

11 Siu, A. L. et al. Screening for depression in adults: US Preventive Services Task Force recommendation statement. JAMA 315, 380–387 (2016). 10.1001/jama.2015.18392

12 ACOG. Screening and Diagnosis of Mental Health Conditions During Pregnancy and Postpartum: ACOG Clinical Practice Guideline No. 4.. Obstetrics & Gynecology 141, 1232–1261 (2023). 10.1097/AOG.0000000000005200

13 Centre of Perinatal Excellence, C. National Perinatal Mental Health Guideline, <https://www.cope.org.au/health-professionals/review-of-new-perinatal-mental-health-guidelines/> (2023).

14 (NIHCE), N. I. f. H. C. E. Antenatal and postnatal mental health: clinical management and service guidance: updated edition. (British Psychological Society 2014).

15 Cox, E. Q., Sowa, N. A., Meltzer-Brody, S. E. & Gaynes, B. N. The perinatal depression treatment cascade: baby steps toward improving outcomes. The Journal of Clinical Psychiatry 77, 20901 (2016). 10.4088/JCP.15r10174

16 Yim, I. S., Tanner Stapleton, L. R., Guardino, C. M., Hahn-Holbrook, J. & Dunkel Schetter, C. Biological and psychosocial predictors of postpartum depression: systematic review and call for integration. Annual Review of Clinical Psychology 11, 99–137 (2015). 10.1146/annurev-clinpsy-101414-020426

17 McCormack, C., Abuaish, S. & Monk, C. Is there an inflammatory profile of perinatal depression? Current Psychiatry Reports 25, 149–164 (2023). 10.1007/s11920-023-01414-y

18 Levin, G. & Ein-Dor, T. A unified model of the biology of peripartum depression. Translational Psychiatry 13, 138 (2023). 10.1038/s41398-023-02439-w

19 Galbally, M., Watson, S. J., Boyce, P., Howard, L. & Herrman, H. Perinatal depression: The use of the Edinburgh Postnatal Depression Scale to derive clinical subtypes. Australian & New Zealand Journal of Psychiatry 58, 37–48 (2024). 10.1177/0004867423119364

20 Hutchens, B. F. & Kearney, J. Risk factors for postpartum depression: an umbrella review. Journal of Midwifery & Women’s Health 65, 96–108 (2020). 10.1111/jmwh.13067

21 Gastaldon, C., Solmi, M., Correll, C. U., Barbui, C. & Schoretsanitis, G. Risk factors of postpartum depression and depressive symptoms: umbrella review of current evidence from systematic reviews and meta-analyses of observational studies. The British Journal of Psychiatry 221, 591–602 (2022). 10.1192/bjp.2021.222

22 Puyané, M. et al. Personality traits as a risk factor for postpartum depression: A systematic review and meta-analysis. Journal of Affective Disorders 298, 577–589 (2022). 10.1016/j.jad.2021.11.010

23 Mohr, D. C., Zhang, M. & Schueller, S. M. Personal sensing: understanding mental health using ubiquitous sensors and machine learning. Annual Review of Clinical Psychology 13, 23–47 (2017). 10.1146/annurev-clinpsy-032816-044949

24 Kamath, J., Barriera, R. L., Jain, N., Keisari, E. & Wang, B. Digital phenotyping in depression diagnostics: integrating psychiatric and engineering perspectives. World Journal of Psychiatry 12, 393 (2022). 10.5498/wjp.v12.i3.393

25 Price, G. D., Heinz, M. V., Song, S. H., Nemesure, M. D. & Jacobson, N. C. Using digital phenotyping to capture depression symptom variability: detecting naturalistic variability in depression symptoms across one year using passively collected wearable movement and sleep data. Translational Psychiatry 13, 381 (2023). 10.1038/s41398-023-02669-y

26 Abd-Alrazaq, A. et al. Systematic review and meta-analysis of performance of wearable artificial intelligence in detecting and predicting depression. NPJ Digital Medicine 6, 84 (2023). 10.1038/s41746-023-00828-5

27 Leaning, I. E. et al. From smartphone data to clinically relevant predictions: A systematic review of digital phenotyping methods in depression. Neuroscience & Biobehavioral Reviews, 105541 (2024). 10.1016/j.neubiorev.2024.105541

28 Krishnamurti, T. et al. Using natural language from a smartphone pregnancy app to identify maternal depression. Res Sq (2023). 10.21203/rs.3.rs-2583296/v1

29 Allen, K. C., Davis, A. & Krishnamurti, T. Indirect identification of perinatal psychosocial risks from natural language. IEEE transactions on affective computing 14, 1506–1519 (2021). 10.1109/TAFFC.2021.3079282

30 Allen, K. et al. Digital phenotyping of depression during pregnancy using self-report data. Journal of Affective Disorders 364, 231–239 (2024). 10.1016/j.jad.2024.08.029

31 Hahn, L. et al. Early identification of postpartum depression using demographic, clinical, and digital phenotyping. Translational Psychiatry 11, 121 (2021). 10.1038/s41398-021-01245-6

32 Zhang, W. et al. The Relationship Between Images Posted by New Mothers on WeChat Moments and Postpartum Depression: Cohort Study. J Med Internet Res 22, e23575 (2020). 10.2196/23575

33 De Choudhury, M., Counts, S., Horvitz, E. J. & Hoff, A. in Proceedings of the 17th ACM conference on Computer supported cooperative work & social computing. 626–638.

34 Fatima, I. et al. Prediction of postpartum depression using machine learning techniques from social media text. Expert Systems 36, e12409 (2019). 10.1111/exsy.12409

35 Suganthi, D. & Geetha, A. Predicting Postpartum Depression with Aid of Social Media Texts Using Optimized Machine Learning Model. International Journal of Intelligent Engineering & Systems 17 (2024). 10.22266/ijies2024.0630.33

36 Hurwitz, E. et al. Harnessing Consumer Wearable Digital Biomarkers for Individualized Recognition of Postpartum Depression Using the All of Us Research Program Data Set: Cross-Sectional Study. JMIR mHealth and uHealth 12, e54622 (2024). 10.2196/54622

37 Slyepchenko, A., Minuzzi, L., Reilly, J. P. & Frey, B. N. Longitudinal changes in sleep, biological rhythms, and light exposure from late pregnancy to postpartum and their impact on peripartum mood and anxiety. The Journal of Clinical Psychiatry 83, 39211 (2022). 10.4088/JCP.21m13991

38 Pitsillos, T. et al. Association between objectively assessed sleep and depressive symptoms during pregnancy and post-partum. Frontiers in Global Women’s Health 2, 807817 (2022). 10.3389/fgwh.2021.807817

39 Micheletti, M. E. Objective markers of mother and infant behavior predict maternal mental health: evidence from a multimodal sensing paradigm, PhD thesis, [The University of Texas at Austin] (2020).

40 Zhong, M., Van Zoest, V., Bilal, A. M., Papadopoulos, F. & Castellano, G. in Proceedings of the 2022 International Conference on Multimodal Interaction. 455–467.

41 Fransson, E. et al. Differentiated mental health patterns in pregnancy during COVID-19 first two waves in Sweden: a mixed methods study using digital phenotyping. Scientific Reports 12, 21253 (2022). 10.1038/s41598-022-25107-3

42 Page, M. J. et al. The PRISMA 2020 statement: an updated guideline for reporting systematic reviews. Systematic Reviews 10, 1–11 (2021). 10.1136/bmj.n71

43 Ouzzani, M., Hammady, H., Fedorowicz, Z. & Elmagarmid, A. Rayyan—a web and mobile app for systematic reviews. Systematic Reviews 5, 1–10 (2016). 10.1186/s13643-016-0384-4

44 Richardson, W. S., Wilson, M. C., Nishikawa, J. & Hayward, R. S. The well-built clinical question: a key to evidence-based decisions. ACP Journal Club 123, A12–13 (1995).

45 Wolff, R. F. et al. PROBAST: a tool to assess the risk of bias and applicability of prediction model studies. Annals of Internal Medicine 170, 51–58 (2019). 10.7326/M18-1376

46 Hummel, A. D. et al. Perinatal depression and its impact on infant outcomes and maternal-nurse SMS communication in a cohort of Kenyan women. BMC Pregnancy and Childbirth 22, 723 (2022). 10.1186/s12884-022-05039-6

47 Hurwitz, E. et al. Unlocking the potential of wearable device wear time to enhance postpartum depression screening and detection. medRxiv (2024). 10.1101/2024.10.07.24315026

48 Tibshirani, R. Regression shrinkage and selection via the lasso. Journal of the Royal Statistical Society Series B: Statistical Methodology 58, 267–288 (1996). 10.1111/j.2517-6161.1996.tb02080.x

49 Currey, D. & Torous, J. Increasing the value of digital phenotyping through reducing missingness: a retrospective review and analysis of prior studies. BMJ Ment Health 26 (2023). 10.1136/bmjment-2023-300718

50 Gabr, M. I., Helmy, Y. M. & Elzanfaly, D. S. Effect of missing data types and imputation methods on supervised classifiers: An evaluation study. Big Data and Cognitive Computing 7, 55 (2023). 10.3390/bdcc7010055

51 Hua, Y. et al. Large language models in mental health care: a scoping review. arXiv preprint arXiv:2401.02984 (2024). 10.48550/arXiv.2401.02984

52 Malgaroli, M., Hull, T. D., Zech, J. M. & Althoff, T. Natural language processing for mental health interventions: a systematic review and research framework. Translational Psychiatry 13, 309 (2023). 10.1038/s41398-023-02592-2

53 Birk, R. H. & Samuel, G. Digital phenotyping for mental health: reviewing the challenges of using data to monitor and predict mental health problems. Current Psychiatry Reports 24, 523–528 (2022). 10.1007/s11920-022-01358-9

