## Supplementary Table 2. Search boolean for "A review of the application of digital phenotyping in predicting peripartum depressive symptoms"

**Supplementary Table 2.** Databases, keywords, and search strategies used in the review

Six databases, PubMed, PsycINFO, CINAHL, Web of Science, CINAHL, Cochrane Trials, and Scopus, were included in the search. In this document, we listed the keywords and the strategy that were implemented. In addition, Google scholar was used as a supplementary tool.

**Keywords**

| **Keyword Group I** | **Keyword Group II** |
| --- | --- |
| Antenatal depression*  Prenatal depression  Postpartum depression  Perinatal depression  Peripartum depression  Maternal depression | Digital phenotyping  Wearable device  Passive digital data  Smartphone  Mobile phone  Mobile application  Real-time data  Ecological momentary assessment  Smart phone application  Text message  Digital behavior  Social media  Online social network  Fitness device |

**Search strategy**

**PubMed search strategy**: “(antenatal depression OR prenatal depression OR postpartum depression OR peripartum depression OR peripartum depression OR maternal depression) AND (digital phenotyping OR wearable device OR passive digital data OR smartphone OR mobile phone OR ecological momentary assessment OR smartphone application OR mobile application OR real-time data OR text message OR digital behavior OR social media OR online social network OR fitness device)” 🡺

Same keywords and Booleans were applied in other databases.

Antenatal depression* or Prenatal depression* or Postpartum depression* or Perinatal depression* or Peripartum depression* or Maternal depression*

Digital phenotyping* or Wearable device* or Passive digital data* or Smartphone* or Mobile phone* or Mobile application* or Real-time data* or Ecological momentary assessment* or Smart phone application* or Text message* or Digital behavior* or Social media* or Online social network* or Fitness device*

[For Scopus]

Antenatal depression* OR Prenatal depression* OR Postpartum depression* OR Perinatal depression* OR Peripartum depression* OR Maternal depression*

Digital phenotyping* OR Wearable device* OR Passive digital data* OR Smartphone* OR Mobile phone* OR Mobile application* OR Real-time data* OR Ecological momentary assessment* OR Smart phone application* OR Text message* OR Digital behavior* OR Social media* OR Online social network* OR Fitness device*
