## Supplementary Table 3. PICO inclusion and exclusion criteria for "A review of the application of digital phenotyping in predicting peripartum depressive symptoms"

**Supplementary Table 3.** Inclusion­–Exclusion criteria for title, abstract, and full article evaluation based on PICO

|  | Inclusion criteria | Exclusion criteria |
| --- | --- | --- |
| **P (population)** | - Women during pregnancy and/or 12 months postpartum | - Women with other psychiatric disorders (e.g., bipolar) - Animal studies |
| **I (indicator)** | - Women with who provided passive digital information (including mobile phone, wearable fitness devices) and/or active digital data, such as mood log, app text messages, etc.) | - Using digital intervention targeting on women with depression. The intervention program contains modules of psychological treatment, cognitive behavioural therapy, etc. - One-time digital data from questionnaires or surveys (without continuous measurement) |
| **C (comparator)** | - (If reported) Active digital data, e.g., self-report for monitoring peripartum depression - Studies combined self-report information or electronic health data related to prediction of peripartum depression | - Studies did not apply any digital devices |
| **O**  **(outcome)** | - Predictive relationships between digital phenotyping patterns and peripartum depression among women during pregnancy and/or 12 months postpartum | - Studies reported only descriptive results or focused on the effect of digital intervention on mothers and/or infants will be excluded. |
| **Others** | - App or devices for pregnancy monitoring or screening purposes (NOT intervention) - Data content can be: location, activity, sleep, social activity, heart rate variability (HRV), screen time, ecological momentary data, etc. - Screening, random control trials (RCT), and observational studies | - Study protocol / cohort profile - Review - Studies focused on the partner only - Feasibility study testing or only evaluating the acceptance of digital apps - Digital media type (TV, radio or other conventional apparatus) |
